# Digital motor and cognitive variability as early markers of the prodromal Parkinson’s disease spectrum

**DOI:** 10.64898/2026.09.06.26358479

**Authors:** H. Chohan, S. Waters, L. Pérez-Carbonell, G. Leschziner, MT. Periñán, B. Huxford, S. Eriksson, JP. Bestwick, A. Schrag, AJ. Noyce, C. Simonet

**Affiliations:** Centre for Preventive Neurology, Wolfson Institute of Population Health, Queen Mary University of London, London, United Kingdom; Sleep Disorders Centre. Guy’s and St Thomas’ NHS Foundation Trust, London, United Kingdom; Department of Clinical and Movement Neuroscience, UCL Institute of Neurology, London, United Kingdom; Department of Neurology, The Royal London Hospital, Barts Health NHS Trust, London, United Kingdom; Department of Neurology, Homerton NHS Foundation Trust, London, United Kingdom; Department of Clinical and Experiential Epilepsy, UCL Institute of Neurology, UCLH/NHS foundation Trust, London, UK; Instituto de Biomedicina de Sevilla (IBiS). Hospital Universitario Virgen del Rocío, Spain

## Abstract

**Background:** Individuals with isolated REM sleep behaviour disorder (iRBD) show motor and cognitive dysfunction prior to clinical α-synucleinopathy. Performance instability may represent an earlier marker of neurodegeneration but remains underexplored using digital tests.

**Objectives:** To assess baseline and longitudinal differences in motor and cognitive performance and examine performance variability in iRBD and related phenotypes.

**Methods:** Three PD-risk-stratified prodromal cohorts (normosmic probable RBD (pRBD), hyposmic pRBD and polysomnography-confirmed iRBD (PSG-iRBD)) were compared with healthy controls. Participants completed the BRadykinesia Akinesia Incoordination (BRAIN) test, Distal Finger Tapping (DFT) test, and Trail Making Test (TMT) remotely. Motor outcomes included kinesia score (KS), akinesia time (AT), and incoordination score (IS); cognitive outcomes included TMT-A/B reaction times and executive function. Baseline differences were analysed using age- and sex-adjusted logistic regression, longitudinal change using linear mixed-effects models, and variability using the mean of squared successive differences (MSSD) and heteroscedastic modelling.

**Results:** At baseline, PSG-iRBD participants showed greater motor and cognitive impairment than controls, hyposmic pRBD demonstrated intermediate deficits and normosmic pRBD participants showed milder but significant impairments. Linear mixed-effects models showed no change in motor trajectories across all groups, while executive function declined significantly in PSG-iRBD (p=0.001). However, variance modelling showed widespread variability. Cognitive variability was elevated in PSG-iRBD and hyposmic pRBD participants (MSSD p<0.035), and heteroscedastic modelling captured significant visit-specific motor instability across all prodromal cohorts on the BRAIN test (all p<0.02).

**Conclusions:** Performance variability precedes measurable linear decline across PD-risk stratified groups. Remote digital testing offers a scalable framework to capture this early instability.

## INTRODUCTION

Isolated REM sleep behaviour disorder (iRBD) and hyposmia (reduced sense of smell) are common prodromal markers of Parkinson’s Disease (PD) and related disorders.^1^ iRBD is a parasomnia characterised by dream-enactment behaviours arising from the loss of normal REM atonia, often resulting in injuries to patients or their bed partners.^2,3^ There is a strong association between iRBD and alpha-synuclein related disorders.^4–6^ Post-mortem studies suggest that up to 94% of people with iRBD show evidence of alpha-synuclein deposition in their brain and greater pathological burden correlates with more severe RBD symptoms.^7^

Most individuals with iRBD eventually develop a neurodegenerative disorder, most commonly PD (44-54%), dementia with Lewy bodies (25-44%) or multiple system atrophy (5-6%).^8,9^ Longitudinal multicentre studies report annual phenoconversion rates of approximately 6%, with cumulative conversion rates exceeding 70% after 12 years of follow-up.^10^ The interval between iRBD diagnosis and overt motor PD is often greater than a decade.^11^ During this prodromal period, subtle abnormalities in quantitative motor performance, gait, handwriting, speed and bradykinesia may emerge several years before diagnosis.^10,12–14^ These findings support the existence of a prolonged prodromal phase characterised by progressive motor dysfunction prior to clinical PD.

Olfactory dysfunction is an early and independent marker of synucleinopathy^15,16^ that affects approximately 70–90% of individuals with PD and often preceding clinical onset by many years.^17^ Meta-analytic evidence shows hyposmia is associated with a 3.84-fold increased risk of developing PD over 3–17 years^18^, indicating it may occur more than a decade before diagnosis. Other prodromal features, including mild parkinsonian signs and cognitive impairment, particularly in attention and executive function, also increase phenoconversion risk. Hyposmia is highly prevalent in RBD, with up to 97% of RBD patients demonstrating increased olfactory thresholds and 63% exhibiting anosmia.^17–22^ However, while iRBD strongly predicts synucleinopathy, many iRBD patients remain isolated for over a decade.

Importantly, the prodromal PD phase is likely to be characterised not only by gradual change in mean performance but also by increasing intra-individual variability, which may precede overt clinical decline.^23–25^ Cross-sectional analyses can miss these fluctuations; therefore longitudinal assessment is essential.

The aim of this study was to explore whether motor and cognitive deficits extend from iRBD to probable RBD (pRBD) with and without hyposmia, to determine whether longitudinal variability in performance can be detected in addition to changes in mean performance over time; and further test remote markers of motor and cognitive assessment.

Longitudinal follow-up of at-risk populations, particularly individuals with iRBD, may improve understanding of prodromal disease trajectories and the interval between iRBD onset and phenoconversion. Such information could help define therapeutic windows for disease-modifying intervention and optimise clinical trial enrichment strategies by identifying individuals at highest risk of conversion^13–15^.

## METHODS

### Study population

Participants defined as having iRBD were identified through the PREDICT-PD study (REC Ref 10/H07/16/85).^17^ PREDICT-PD is an online cohort study which aims to identify populations with an increased risk of PD. iRBD participants from the PREDICT-PD study were recruited from the Sleep Clinic at Guy’s and St Thomas’ NHS Foundation Trust and University College London Hospital. They all had a video-polysomnography performed following American Academy of Sleep Medicine standards^27^, and their RBD diagnosed as per current International Classification of Sleep Disorders criteria.^28^ Questionnaire-based measures of dream enactment behaviour may identify a broader and potentially earlier prodromal population than PSG-RBD, despite reduced specificity.^18^ Participants with pRBD and controls were identified through the PREDICT-PD study.

To enrich for underlying synucleinopathy risk, this group was further stratified according to the presence of hyposmia. Individuals with pRBD were defined by answering *yes* responses to the single item RBD questionnaire (RBD1Q; “Have you ever been told, or suspected yourself, that you seem to ‘act out your dreams’ while asleep (for example, punching, flailing your arms in the air, making running movements etc.?”).^18^ Participants with pRBD who scored within the worst age- and sex-specific 15th centile on either the 40-item or 6-item UPSIT were classified as pRBD with hyposmia.

Exclusion criteria included <50 years old, a diagnosis of a neurodegenerative disease (including PD, Dementia with Lewy bodies, Multiple Sclerosis, Dementia, and Motor Neurone Disease), restless legs syndrome, tremor, stroke and anti-psychotic drug use. The PREDICT-PD study has devised a risk algorithm which considers intermediate risk factors such as tapping speed, hyposmia and RBD as markers of prodromal PD. The algorithm includes intermediate markers into the risk model.^17,19^. Therefore, any controls classified as ‘high-risk’ via the PREDICT-PD risk algorithm^17,19^ were removed from the analysis. Controls not identified as ‘high-risk’ were included in this study. Figure 1 shows the recruitment process for this study along with the cohort size per group and analyses.

**Figure 1.**
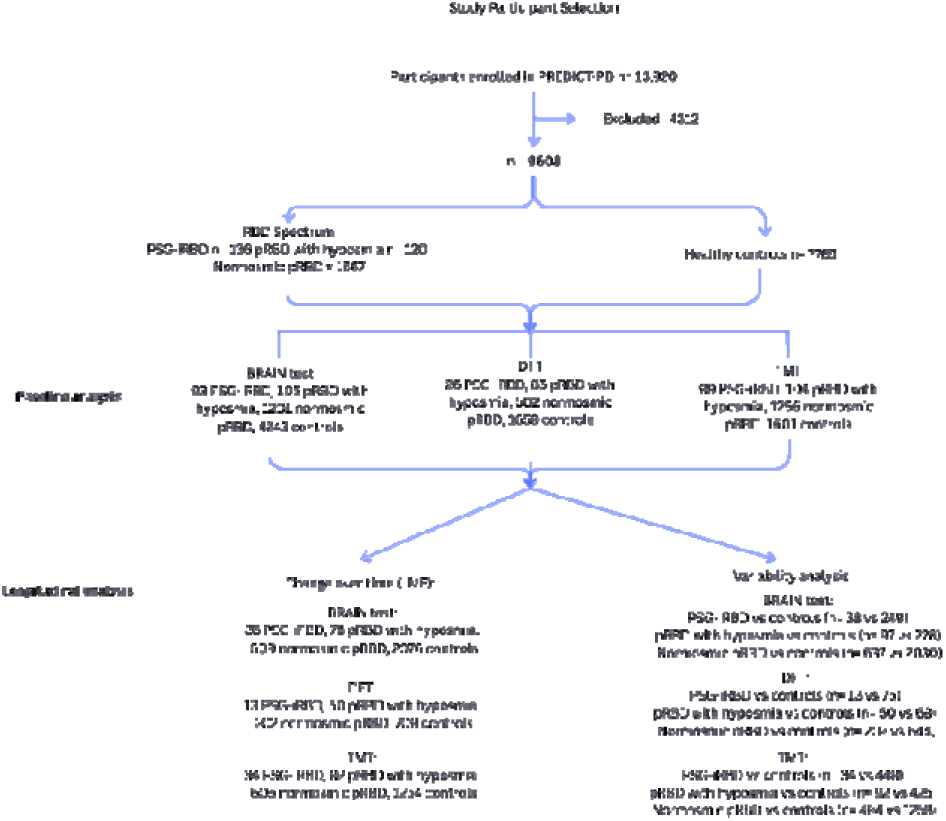
Flow diagram of participant selection and inclusion in baseline and longitudinal digital motor-cognitive analyses. Participants were recruited from the PREDICT-PD cohort and categorised into PSG-iRBD, pRBD with hyposmia, normosmic pRBD and healthy controls. The diagram outlines the numbers of participants included in for the BRAIN test, Distal Finger Tapping test (DFT), and Trail Making Test (TMT), and those contributing to longitudinal mixed-effects models and variability analyses. pRBD - probable REM sleep behaviour disorder; PSG-iRBD - polysomnography confirmed isolated REM sleep behaviour disorder; LME - linear mixed effects model.

### Motor assessment

The BRadykinesia Akinesia Incoordination (BRAIN) test is a 30-second web-based alternating finger keyboard tapping test validated for motor dysfunction in PD and iRBD.^20,21,29^ The Distal Finger Tapping Test (DFT) remotely tests distal index repetitive finger tapping movement over 20 seconds and is similarly validated.^30^ Kinetic parameters for both tests include the kinesia score (KS, number of key taps over time (20s/30s)), akinesia time (AT, mean dwell-time on each key) and incoordination score (IS, variance of travelling time between key taps).

### Cognitive assessment

Cognitive function was assessed using the Trail Making Test (TMT).^31,32^ The TMT assesses visual attention, processing speed, mental flexibility and executive function.^32^ In TMT-A, participants sequentially connect 25 encircled numbers in numerical order, whilst TMT-B requires alternating numbers and letters. The TMT provided reaction times and executive function scores. Executive function scores were derived by subtracting TMT-B reaction time from TMT-A reaction time.

### Statistical analysis

Parameters from the keyboard tapping tasks and TMT scores were analysed cross-sectionally and longitudinally. Normality was assessed visually (density and Q-Q plots) and with Shapiro-Wilk tests; non-normal scores were log-transformed. Baseline demographics and clinical data were compared using ANOVA for continuous and chi-squared/Fisher’s exact tests for categorical variables (α = 0.05). Binary logistic regression models used group status as the outcome and z-standardised test scores as predictors, with KS direction reversed so higher values reflected greater impairment. Odds ratios reflect the change in odds of group membership per 1 SD worsening in performance. Age² accounted for nonlinear age effects.

Model fit was checked via Hosmer-Lemeshow tests. Worst-hand scores maximised sensitivity to subtle unilateral impairment, and control data were winsorised to mitigate extreme values likely reflecting artefacts rather than pathology. TMT models were additionally adjusted for age, sex and education level.

Keyboard tapping composites were derived using longitudinal confirmatory factor analysis (CFA), with a single latent factor at each timepoint defined by KS, IS and AT (z-standardised). Model fits were assessed using the comparative fit index (CFI), Tucker-Lewis index (TLI), root mean square error of approximation (RMSEA) and standardised root mean square residual (SRMR). Lacking full metric invariance, partial longitudinal invariance was applied, and factor scores were extracted via full information maximum likelihood. The TMT lacked a stable latent structure; therefore, a composite was calculated from z-standardised Trail B and executive function (Trail B − Trail A). A combined composite was used for exploratory analyses.

Linear mixed-effects models assessed changes in each kinetic parameter over time and group-by-time interactions, adjusting for age (including a quadratic term) and sex. Models included fixed effects for time, group, and their interaction, with participant-level random intercepts for repeated measures. Likelihood ratio tests guided model selection for nested models. Random slopes were not included due to many observations per person over time. Participants completed a median of 2 sessions (range 1–3) approximately 1 year apart, and analyses included those with baseline and at least one follow-up assessment to assess within-person change. Mean follow-up across the tests was 2.03 years.

Variance analyses assessed motor and cognitive score changes over time. Cases and controls were balanced on age and sex using nearest-neighbour propensity score matching (1:5 matching), with a 0.2 SD caliper for age and exact matching for sex. Group-level dispersion was summarised with means, SDs, and bootstrapped confidence intervals, with outcome values winsorised at the 1st and 99th percentiles. Intra-individual variability was characterised using subject-level SD and mean square of successive differences (MSSD) and compared via Wilcoxon rank-sum tests. Variability was further assessed using heteroscedastic mixed-effects models (varIdent) which allowed residual variance to differ across visits. These models were compared with homoscedastic equivalents using likelihood ratio tests and AIC/BIC.

All analyses were performed in R (version 4.5.1) and the code can be found on GitHub (https://github.com/Wolfson-PNU-QMUL/motor_cognition_iRBD). Cases and controls were matched using the MatchIt package. Logistic regression models were fitted using glm(), CFA was conducted using lavaan, linear mixed-effects models were fitted using lme4 and lmerTest, and heteroscedastic mixed-effects models were fitted using nlme. Statistical significance was defined as p<0.05. As analyses were exploratory, no formal correction for multiple comparisons was applied.

## RESULTS

Groups were age-matched. Cases (PSG-iRBD, hyposmic pRBD, and normosmic pRBD) were predominantly male, whereas controls were mainly female (all p<0.05). Full demographic and baseline performance data are presented in Supplementary Tables S1-S2 and Figure S1. Cross-sectional and longitudinal model estimates are provided in Supplementary Table S3 and S5, respectively, and mean changes and trajectories are shown in Supplementary Table S4 and Figure S2.

### Cross-sectional analysis at baseline

#### BRAIN test

At baseline, PSG-iRBD participants showed significantly worse BRAIN test digital motor measures compared with controls (Figure 2, Table S3). The strongest association was observed for KS (OR: 2.16, 95% CI: 1.67 to 2.81, DoF: 4331, p<0.001) followed by AT (OR: 1.55, 95% CI: 1.27 to 1.89, DoF: 4331, p<0.001) and IS (OR 1.25, 95% CI: 1.01 to 1.55, DoF: 4331, p=0.043). Similarly, hyposmic pRBD participants also showed poorer performance compared to controls, although effect sizes were smaller for KS (OR 1.60, 95% CI: 1.28 to 2.02, p<0.001). Normosmic pRBD participants performed significantly worse than controls across all BRAIN test measures (all p<0.001).

**Figure 2.**
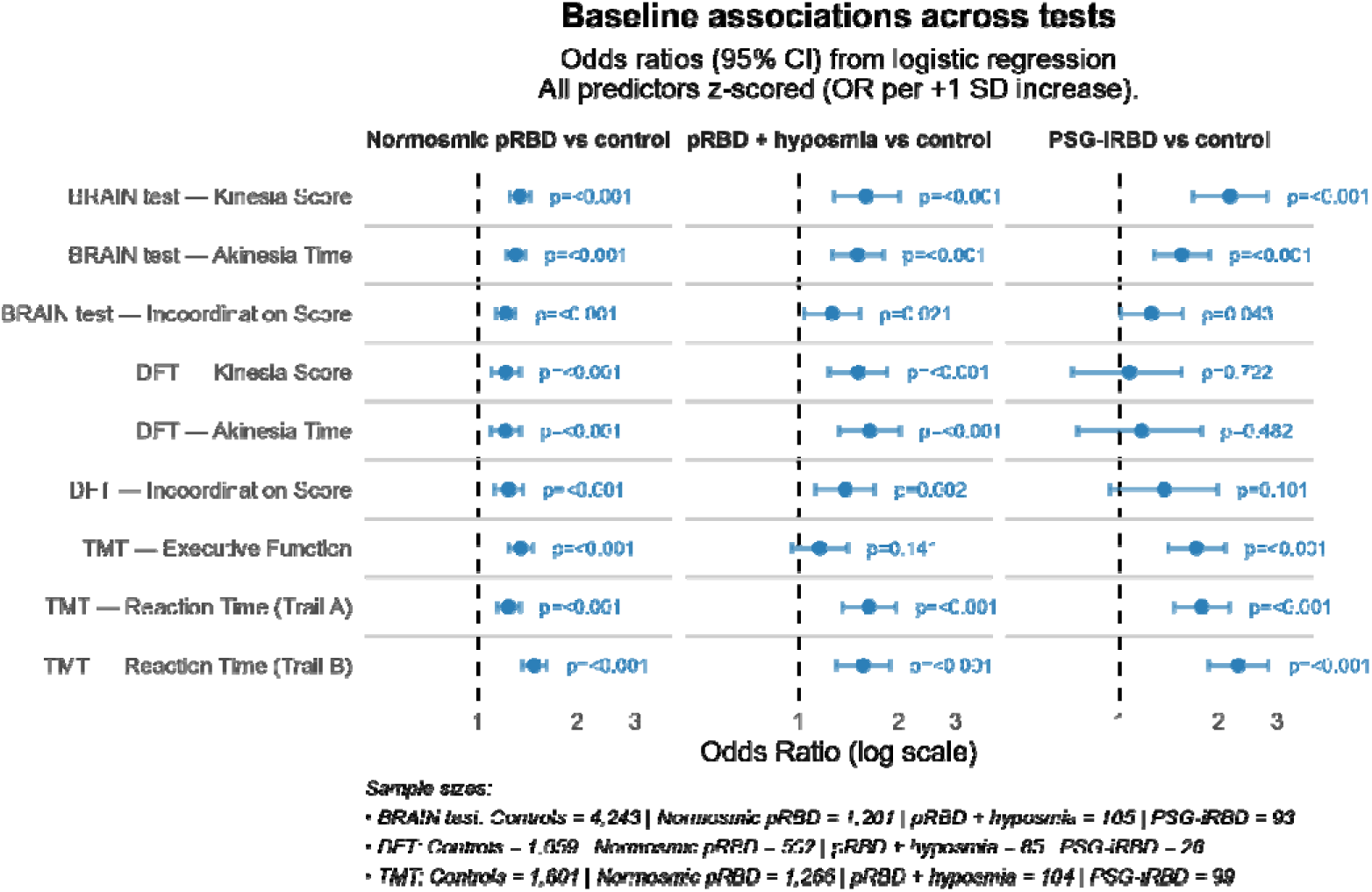
Forest plot show baseline test performance. Forest plot of adjusted differences in digital motor-cognitive performance between prodromal PD groups and controls. Results demonstrate widespread baseline impairments across motor and cognitive domains in groups compared with controls. pRBD - probable REM sleep behaviour disorder, PSG-iRBD - polysomnography confirmed isolated REM sleep behaviour disorder, BRAIN test - Bradykinesia Akinesia Incoordination test, DFT - Distal Finger Tapping Test, TMT - Trail Making Test.

#### DFT

With respect to DFT, PSG-iRBD participants tended to perform worse than controls across KS, AT, and IS, though differences were not statistically significant (IS OR 1.37, 95% CI: 0.93 to 1.98, DoF: 1680, p=0.101; Table S3). In contrast, hyposmic pRBD participants performed significantly worse across all measures, with the strongest effect for AT (OR 1.64, 95% CI: 1.33 to 2.03, DoF: 1740, p<0.001). Normosmic pRBD participants also showed significant but smaller impairments across parameters (e.g., IS OR 1.24, 95% CI 1.12 to 1.37, DoF: 2156, p<0.001) (Figure 2; Table S3).

#### TMT

PSG-iRBD participants showed significantly worse cognitive performance on the TMT than controls, with slower completion times for both TMT-A and TMT-B and greater executive dysfunction (executive function OR 1.71, 95% CI 1.40 to 2.09, DoF: 1694, p<0.001; Table S3). Hyposmic pRBD participants also performed slower on TMT-A and TMT-B (TMT-A reaction time OR 1.63, DoF: 1700, 95% CI 1.36 to 1.97, p<0.001), although executive function did not differ significantly. Furthermore, normosmic pRBD participants performed worse than controls across all TMT measures, albeit with smaller effect sizes (executive function OR 1.35, 95% CI: 1.24 to 1.46, DoF: 2861, p<0.001) (Figure 2; Table S3).

### Longitudinal trajectories, variability and composite measures

#### Composite scores

The combined motor-cognitive composite showed a significant group-by-time interaction (F=20.29, p<0.001). Relative to controls, composite scores declined more rapidly in PSG-iRBD (β = -0.11, 95% CI -0.14 to -0.07, p<0.001) and normosmic pRBD participants (β=-0.024, 95% CI -0.03 to -0.01, p<0.001), whereas no significant difference in the rate of decline was observed in hyposmic pRBD participants (β=0.023, 95% CI -0.01 to 0.05, p=0.105).

#### BRAIN test

Over time, longitudinal mixed-effects models did not identify significant group-by-time interactions for BRAIN test measures in PSG-iRBD participants compared with controls (KS β=- 0.01, 95% CI -2.13 to 2.12, t =-0.01, p=0.996; Table S5). Normosmic pRBD participants also showed no differences in longitudinal trajectories across BRAIN parameters (Table S5). In hyposmic pRBD participants, a trend towards worsening AT over time was observed compared with controls (β=3.44, 95% CI -0.74 to 7.17, t=1.81, p=0.070), although this did not reach statistical significance.

Variability analyses demonstrated evidence of increased motor instability across prodromal PD groups. In PSG-iRBD participants, within-subject variability in IS did not differ significantly from controls using MSSD (p=0.134); however, heteroscedastic modelling demonstrated significantly greater visit-specific residual variance (LRT=30.72, p<0.001), corresponding to approximately 45% higher variability at intermediate visits (Figure 3). AT and KS showed a similar pattern, with no differences detected using MSSD (p=0.680 and p=0.395) but significant visit-specific heteroscedasticity (LRT=12.36, p=0.006 and LRT=9.90, p=0.019). In hyposmic pRBD participants, MSSD did not detect variability differences for IS, AT or KS compared with controls (p=0.230, 0.816 and 0.711, respectively); however, heteroscedastic mixed-effects models revealed significant visit-specific residual variance across all measures (IS LRT=101.90, AT LRT=29.39, KS LRT=22.22; all p<0.001). By comparison, normosmic pRBD participants showed greater within-subject variability than controls for IS and AT using MSSD (p=0.013 and p=0.017), but not KS (p=0.062). Heteroscedastic modelling likewise demonstrated significant visit-specific variance across all parameters (IS LRT=371.40, AT LRT=99.62, KS LRT=18.11; all p<0.001).

**Figure 3.**
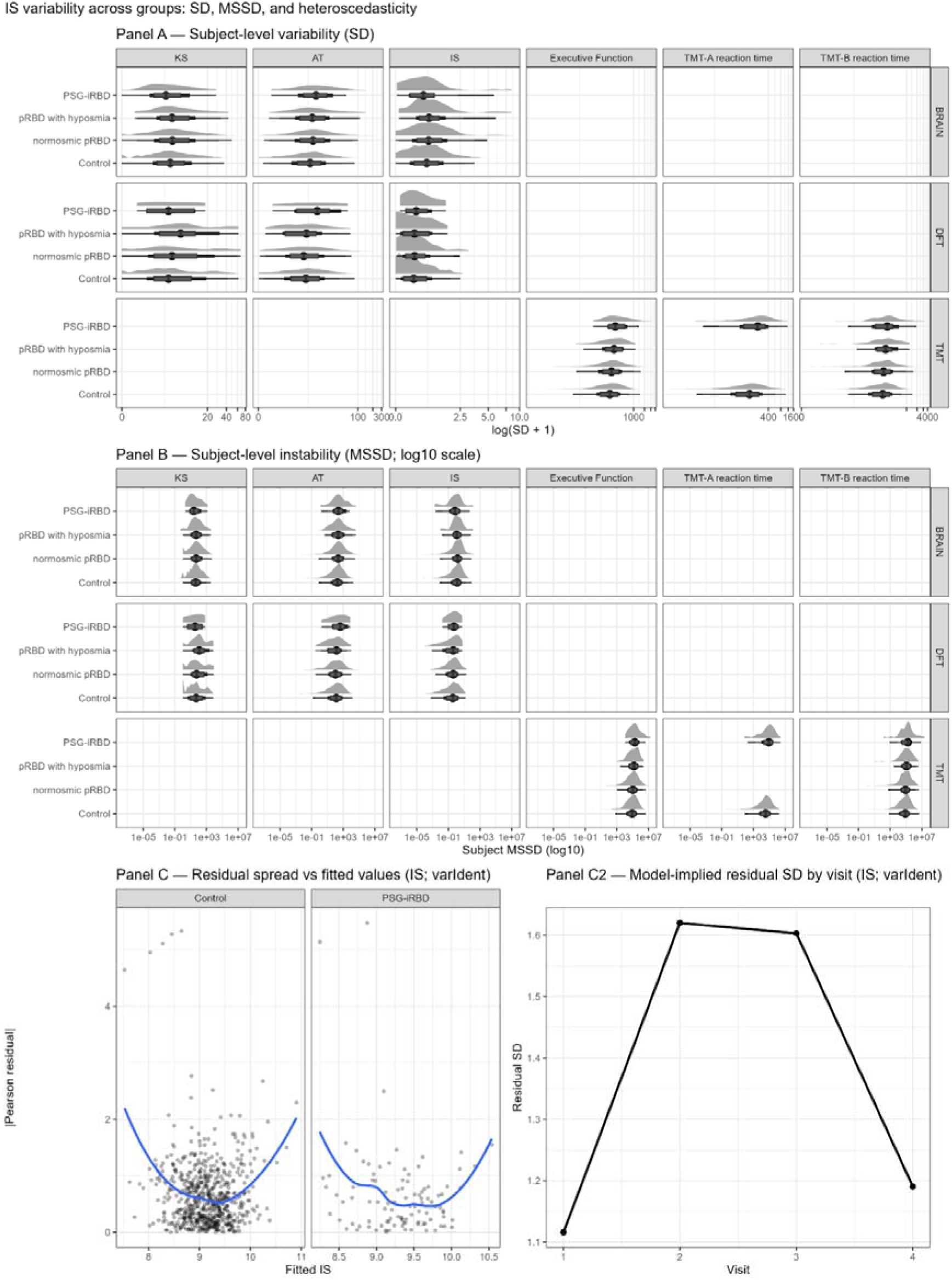
Figure 3 Variability and instability in digital motor-cognitive performance across prodromal Parkinson’s disease groups. Panel A shows subject-level variability measured using the standard deviation (SD) across repeated assessments. Panel B shows within-subject instability quantified using the mean squared successive difference (MSSD; log10 scale). Panel C illustrates heteroscedasticity in the mixed-effects models, including residual spread versus fitted values (C) and model-implied residual standard deviation across study visits (C2). Outcomes are derived from the BRAIN test IS. IS - incoordination score, BRAIN test - Bradykinesia Akinesia Incoordination test, DFT - Distal Finger Tapping Test, TMT - Trail Making Test.

#### DFT

Across all prodromal groups, longitudinal mixed-effects models did not show significant group-by-time interactions for any DFT measure across prodromal groups compared with controls (Table S5). In PSG-iRBD participants, IS and KS trajectories did not differ from controls (IS: β=- 0.25, 95% CI -1.39 to 0.89, t=-0.43, p=0.664). Although AT trajectories differed numerically, this was not significant (β=-19.4, 95% CI -56.8 to 18.0, t=-1.02, p=0.310). Similarly, no significant longitudinal differences were observed in pRBD participants with hyposmia or normosmic pRBD across IS, AT or KS (Table S5).

Variability analyses did not reveal group differences. Within-subject variability in KS, AT and IS did not differ between PSG-iRBD participants and controls using MSSD (all p≥0.518), and heteroscedastic models did not improve model fit (all LRT p≥0.091). Similarly, no variability or visit-specific heteroscedasticity differences were observed in pRBD participants with hyposmia or normosmic pRBD participants (all MSSD and LRT p≥0.085).

#### TMT

Longitudinal analyses indicated worsening executive function in PSG-iRBD participants compared to controls (β=-130.0, 95% CI -203.0 to -56.3, t=-3.47, p=0.001, Table S5). Reaction times for TMT-A and TMT-B showed little change relative to controls, although a modest difference in TMT-B trajectories was observed (β=0.32, 95% CI 0.05 to 0.59, t=2.32, p=0.020). In contrast, no significant group-by-time interactions were observed in pRBD participants with hyposmia or normosmic pRBD participants for executive function or reaction time measures (Table S5).

Variability analyses revealed greater within-subject fluctuations in cognitive performance across prodromal PD groups. PSG-iRBD participants showed greater variability than controls for TMT-A reaction time and executive function using MSSD (p=0.015 and p=0.020), although no difference was observed for TMT-B. Heteroscedastic mixed-effects models indicated significant visit-specific variance for TMT-B and executive function (both LRT p<0.001). Hyposmic pRBD participants exhibited greater variability for TMT-B reaction time and executive function compared with controls (MSSD p=0.034 and p=0.010), while TMT-A variability did not differ significantly. Heteroscedastic modelling similarly demonstrated visit-specific variance for TMT-B and executive function (both LRT p<0.001). In normosmic pRBD participants, no group differences in variability were detected using MSSD; however, heteroscedastic models indicated significant visit-specific variance across all TMT measures (all LRT p<0.001).

## DISCUSSION

Using remote, quantitative tests, this study shows that PD-risk-stratified prodromal cohorts show motor and cognitive deficits and increased variability. While mean longitudinal trajectories of motor and cognitive function appear stable, significant visit-to-visit performance fluctuations are seen through heteroscedastic modeling and are widespread across all prodromal cohorts included in this study. PSG-iRBD participants showed the greatest deficit and instability, while hyposmic pRBD participants demonstrated an intermediate phenotype with clear baseline impairments and significant longitudinal variability. Conversely, normosmic pRBD participants demonstrated subtle mean differences with evidence of visit-specific fluctuations over time. This indicates heterogeneity within normosmic pRBD populations rather than a simple earlier prodromal stage. Preserved olfaction may signal alternative synucleinopathy phenotypes, including multiple system atrophy, although definitive longitudinal phenoconversion data are required to clarify these trajectories.^33^ Overall, measuring variability can improve risk stratification, identify high-risk individuals for clinical trials and advance digital biomarker development.

Olfactory dysfunction strongly correlates with increased phenoconversion risk, greater underlying synuclein pathology and SAA positivity in iRBD.^34^ Hyposmic iRBD participants show greater motor and cognitive dysfunction and are more likely to develop synucleinopathy.^35,36^ In our study, pRBD participants with hyposmia showed an intermediate phenotype, performing worse than normosmic pRBD participants but less impaired than PSG-iRBD. This supports existing research implying that hyposmia reflects a greater disease burden and phenoconversion likelihood.^15,37–39^ Importantly, our findings show how stratifying pRBD by smell function is a meaningful endeavour not just for baseline-risk but also for capturing the underlying dynamics of disease progression. Although hyposmic pRBD participants showed relatively stable longitudinal trajectories, heteroscedastic modelling revealed significant visit-specific residual variance across all BRAIN and TMT measures (all LRT p<0.001). This indicates that the combination of hyposmia, an α-synucleinopathy and prodromal symptoms manifests as instability rather than a linear decline. Within this hyposmic cohort, a selective variability pattern emerged. BRAIN test fluctuations occurred without consistent MSSD increases, suggesting an intermittent motor instability. However, cognitive variability was more pronounced, with greater TMT-B and executive function fluctuations relative to controls. Thus, pRBD with hyposmia may reflect a more advanced prodromal stage where instability is primarily detectable in executive function rather than across motor performance.

Longitudinal analyses used a CFA-derived composite score, with partial invariance supporting comparability across time. Group-by-time interactions were generally absent for finger-tapping measures, suggesting relative stability in average performance during the prodromal phase.

While this seems to contradict evidence that quantitative motor tests like alternating finger tapping strongly predict incident PD in iRBD cohorts^10^, it highlights a clear methodological nuance. Alternate finger tapping distinguishes healthy controls from iRBD patients cross-sectionally and future converters from non-convertors. However, our lack of longitudinal interaction indicates that the mean rate of change during this specific prodromal window is relatively stable.

In contrast, executive function worsened significantly over time in PSG-iRBD. Prior research using motion-capture tapping protocols (Slow Motion Analysis of Repetitive Tapping (SMART), BRAIN, and DFT) has shown slower, irregular, and less rhythmically consistent tapping in iRBD compared to controls, reflecting early motor instability.^40^ Importantly, this increased inconsistency and variability in tapping with stable mean performance has been proposed as a prodromal marker of PD,^40,41^ consistent with early basal ganglia dysfunction.^42^ This , highlights movement inconsistency, not just slowness, as an early marker of emerging nigrostriatal degeneration.

Participants with iRBD and motor impairment are approximately three times more likely to phenoconvert to an α-synucleinopathy (HR 3.16).^43,44^ The motor deficits observed are consistent with early dopaminergic dysfunction within the basal ganglia.^45^ At baseline, PSG-iRBD participants performed the BRAIN test more arrhythmically than controls and tapped fewer alternating keys, indicating impaired motor timing and execution stability.

Cognitive impairment is a key prodromal feature of PD, with changes in global recognition and executive function detectable up to 6 years before the clinical diagnosis of PD.^46^ Also, cognitive impairment is acknowledged as an independent prodromal marker by the Movement Disorder Society.^26^ Importantly, iRBD is strongly associated with Lewy body disease^47^ and is a core feature for dementia with Lewy bodies (DLB).^48,49^ Previous studies report deficits in processing speed, executive function and cognitive flexibility in iRBD and found worse TMT-B performance in iRBD compared to controls.^50^ Similarly, in our study, PSG-iRBD participants performed the TMT slower than controls and with greater executive dysfunction, showing significant deterioration over time. PSG-iRBD participants also showed slower TMT-A and TMT-B performance and executive dysfunction relative to controls, which worsened over time, indicating frontal-striatal loops supporting cognitive flexibility and executive control in PSG-iRBD.^51^ Overall, these findings indicate that motor and cognitive networks are impaired in PSG- iRBD, consistent with synuclein-related neurodegeneration.

While group differences in motor and cognitive performance provide clear evidence of impairment in iRBD, our variance analysis reveals an additional dimension of dysfunction^52^. Fluctuating visit-to-visit performance suggests that longitudinal design may be a more sensitive prodromal marker. In PSG-iRBD participants, this variability was most pronounced in TMT executive function and TMT-B reaction time, perhaps reflecting early instability in the frontostriatal networks mediating motor timing and cognitive control. Our data shows the value of using two approaches to analyse variability. The MSSD captures rapid sequential fluctuations whilst heteroscedastic modelling investigates how much an individual’s performance deviates from the group average at specific timepoints. Our study revealed highly significant visit-specific heteroscedasticity (e.g. LRT=101.90 in hyposmic pRBD) despite non-significant MSSD changes. This demonstrates that prodromal progression may not be characterised by continuous rapid fluctuation but rather by distinct phases where performance becomes unstable and fluctuates heavily around the group average.

Furthermore, the digital remote tests used in this study capture varying degrees of variability. Normosmic pRBD participants showed fluctuations across visits in BRAIN test performance, despite milder mean impairments relative to those observed in PSG-iRBD, while DFT measures remained largely stable. The lack of variability over time in DFT measures implies that digital motor tools vary in their sensitivity to instability in the early prodromal phase. The TMT was sensitive for executive function, while the BRAIN test captured greater variability over time than the DFT. This is consistent with evidence that proximal bradykinesia measures detect early motor dysfunction more effectively than distal finger movement tests^30,40^ which emphasises the need for multi-model tracking in smell-stratified cohorts. Overall, our results support a continuum of impairment across the RBD spectrum: normosmic pRBD exhibited subtle fluctuations without consistent increase in variability compared with controls and hyposmic pRBD demonstrated greater cognitive variability, more closely resembling the variability observed in PSG-iRBD. Thus, these findings reinforce the idea that hyposmia increases the risk of phenoconversion, and emerging instability may be a more sensitive indicator of prodromal synucleinopathy.

Key strengths of this study include the large PREDICT-PD dataset, the use of mixed-model variance analysis and remote unsupervised testing. While unsupervised assessments introduce environmental noise, they minimise experimenter effects and capture authentic, real-world performance variability. Limitations include inconsistent sample sizes, differences in sex distribution, incomplete follow-up and reliance on observational data. Methodologically, the DFT analyses were limited by small sample sizes and only two timepoints, and motor assessments used worst-hand scores without accounting for handedness. Future studies should incorporate definitive phenoconversion outcomes and blood biomarkers to validate these trajectories. Predictive accuracy could be further enhanced through longer follow-up periods and machine learning risk modelling.

In conclusion, variability appears to differentiate risk groups across the RBD spectrum. Performance variability may precede linear decline, serving as a promising early marker and outcome measure for clinical trials. These findings highlight the potential of remote digital tests for screening and longitudinal monitoring in confirmed and suspected iRBD.

## Data Availability

All data produced in the present study are available upon reasonable request to the authors

## DECLARATIONS

### Availability of Data and Materials

All data analysed and generated in this study are included in this published article and its supplementary information files. The code used for analysis can be found on GitHub.

## Acknowledgments

The PREDICT-PD study is funded by Parkinson’s UK. The Centre for Preventive Neurology is funded by the Barts Charity.

## Author’s Roles

HC, SW, AJN and CS contributed to the conception and design of the study. HC, and SW acquired and analysed the data, and drafted the figures. HC, SW, MTP, AJN and CS contributed to data analysis. HC and CS drafted the manuscript. SW, LPC, GL, MTP, BH, SE, JPB, AS and AJN edited the manuscript.

## Financial Disclosures from all authors

Harneek Chohan: No financial disclosure or conflict of interest to declare.

Sheena Waters: Reports grants from Michael J. Fox Foundation and Alzheimer’s Research UK. Is a co-lead of Imaging Working Group of DEMON (Deep Dementia Phenotyping) Network.

Laura Pérez-Carbonell: Reports grants from Michael J. Fox Foundation. Has consulting fees with Takeda UK and Idorisa and speaker fees for Guy’s and St Thomas’ NHS Foundation Trust.

Guy Leschziner: No financial disclosure or conflict of interest to declare.

Maria Teresa Periñán: Reports grants from Michael J. Fox Foundation.

Brook Huxford: No financial disclosure or conflict of interest to declare.

Sofia Eriksson: Reports honoraria by Angelini Pharma. Is a President elect and chair of EDI committee for the Association of British Neurologists.

Jonathan P Bestwick: No financial disclosure or conflict of interest to declare.

Anette Schrag: Reports fundings from European Commission, Parkinson’s UK, International Parkinson’s and Movement Disorders Society, National Institute of Health (NIHR) and UCLH Biomedical Research Centre. Has royalites of licences with Oxford University Press and University College London. Receives consulting fees from Bial, AbbVie, Boehringer and Merz. On the board of MDS-ES.

Alastair J Noyce: Reports funding from Parkinson’s UK, Barts Charity, Cure Parkinson’s, UKRI, HORIZON 2020, National Institute for Health and Care Research, Innovate UK, Solvemed, the Medical College of Saint Bartholomew’s Hospital Trust, Alchemab, Aligning Science Across Parkinson’s Global Parkinson’s Genetics Program (ASAP-GP2), Michael J. Fox Foundation, Tower Hamlets Council and QMUL Impact Fund. Receives consulting feed from AbbVie, Bial and Roche. Receives payment or honoraria from International Parkinson and Movement Disorders Society. On the board for COBALT clinical trial.

Cristina Simonet: No financial disclosure or conflict of interest to declare.

