## Supplementary Material for "Digital motor and cognitive variability as early markers of the prodromal Parkinson’s disease spectrum"

**Supplementary table S1: Baseline characteristics by group**

|  | Controls | Normosmic pRBD | pRBD with hyposmia | PSG-iRBD |
| --- | --- | --- | --- | --- |
| N | 4340 | 1301 | 106 | 106 |
| Age (mean ± SD) | 67.3 ± 5.1 | 67.8 ± 5.9 | 69.1 ± 5.6 | 68.5 ± 6.7 |
| Sex (% male) | 31.2% | 55.3% | 62.3% | 73.6% |
| Education (% secondary) | 14.8% | 18.1% | 16.0% | 21.7% |
| Smell test score | 4.8 ± 0.9 | 4.7 ± 0.9 | 2.5 ± 1.0 | 3.0 ± 1.6 |

Baseline characteristics of groups. Values are presented as mean ± standard deviation (SD) for continuous variables and percentages for categorical variables. Groups include healthy controls, normosmic pRBD, pRBD with hyposmia, and PSG-iRBD). Smell test scores reflect olfactory performance on the 6-item smell test.

pRBD - probable REM sleep behaviour disorder, PSG-iRBD - polysomnography confirmed isolated REM sleep behaviour disorder.

**Supplementary table S2: Motor and cognitive scores at baseline**

|  | Controls | Normosmic pRBD | pRBD with hyposmia | PSG-iRBD |
| --- | --- | --- | --- | --- |
| BRAIN test | | | | |
| N | 4243 | 1201 | 105 | 93 |
| KS (mean ± SD) | 55.69 ± 13.87 | 50.82 ± 15.22 | 49.52 ± 12.53 | 46.76 ± 12.26 |
| AT (mean ± SD) | 103.49 ± 29.87 | 110.25 ± 40.25 | 116.20 ± 37.83 | 116.53 ± 38.67 |
| IS (median ± IQR) [log-transformed] | 2.22 (2.10–2.31) | 9.57 (8.48–10.50) | 9.61 (8.77–10.34) | 9.60 (8.71–10.43) |
| DFT | | | | |
| N | 1659 | 502 | 85 | 26 |
| KS (mean ± SD) | 76.75 ± 25.77 | 75.55 ± 27.03 | 71.26 ± 26.88 | 81.00 ± 23.54 |
| AT (mean ± SD) | 112.51 ± 36.37 | 115.07 ± 37.37 | 124.92 ± 43.29 | 107.77 ± 40.82 |
| IS (median ± IQR) [log-transformed] | 7.23 (6.52–7.95) | 7.37 (6.74–8.16) | 7.64 (7.03–8.28) | 7.77 (6.99–8.21) |
| TMT | | | | |
| N | 1601 | 1266 | 104 | 99 |
| Executive function (mean ± SD) | 640.03 ± 469.29 | 791.30 ± 570.22 | 723.70 ± 529.62 | 969.31 ± 706.24 |
| TMT-A reaction time (mean ± SD) | 1153.95 ± 370.98 | 1228.93 ± 382.06 | 1354.31 ± 449.37 | 1439.39 ± 575.61 |
| TMT-B reaction time (mean ± SD) | 1793.97 ± 560.00 | 2020.23 ± 686.46 | 2078.01 ± 638.62 | 2408.70 ± 989.61 |

Values are presented as mean ± SD for normally distributed variables and median with IQR for non-normally distributed variables. Measures are derived from the BRAIN test, DFT, and TMT. Groups include healthy controls, normosmic pRBD, pRBD with hyposmia, and PSG-iRBD.

KS = kinesia score; AT = akinesia time; IS = incoordination score; BRAIN test = Bradykinesia Akinesia Incoordination test; DFT = Distal Finger Tapping test; TMT = Trail Making Test; pRBD = probable REM sleep behaviour disorder; PSG-iRBD = polysomnography-confirmed isolated REM sleep behaviour disorder.

**Supplementary Table S3 — Full baseline model outputs from multivariate logistic regression analysis**

| Test | Outcome | Comparison | Covariate | OR (95 % CI) | p value |
| --- | --- | --- | --- | --- | --- |
| BRAIN test | KS | PSG-iRBD vs controls  (93 vs 4243) | Group | 2.16 (1.67 - 2.81) | <0.001 |
|  |  |  | Age^2^ | 1.01 (1.01 - 1.01) | <0.001 |
|  |  |  | Sex (Male) | 5.36 (3.40 - 8.74) | <0.001 |
|  |  | pRBD with hyposmia vs controls  (105 vs 4243) | Group | 1.60 (1.28 - 2.02) | <0.001 |
|  |  |  | Age | 1.06 (1.02 - 1.10) | 0.002 |
|  |  |  | Sex (Male) | 3.15 (2.11 - 4.76) | <0.001 |
|  |  | Normosmic pRBD vs controls  (1201 vs 4243) | Group | 1.34 (1.25 – 1.44) | <0.001 |
|  |  |  | Age^2^ | 1.01 (1.01 - 1.01) | <0.001 |
|  |  |  | Sex (Male) | 2.50 (1.25 - 1.44) | <0.001 |
|  | AT | PSG-iRBD vs controls  (93 vs 4243) | Group | 1.55 (1.27 – 1.88) | <0.001 |
|  |  |  | Age^2^ | 1.01 (1.01 - 1.01) | <0.001 |
|  |  |  | Sex (Male) | 7.09 (4.49 - 11.59 | <0.001 |
|  |  | pRBD with hyposmia vs controls  (105 vs 4243) | Group | 1.51 (1.26 -1.81) | <0.001 |
|  |  |  | Age | 1.06 (1.02 - 1.10) | 0.002 |
|  |  |  | Sex (Male) | 3.97 (2.66 - 5.99) | <0.001 |
|  |  | Normosmic pRBD vs controls  (1201 vs 4243) | Group | 1.30 (1.22 – 1.39) | <0.001 |
|  |  |  | Age^2^ | 1.01 (1.01 - 1.01) | <0.001 |
|  |  |  | Sex (Male) | 2.87 (2.51 - 3.28) | <0.001 |
|  | IS | PSG-iRBD vs controls  (93 vs 4243) | Group | 1.25 (1.01 - 1.55) | 0.043 |
|  |  |  | Age^2^ | 1.01 (1.01 - 1.01) | <0.001 |
|  |  |  | Sex (Male) | 6.19 (3.93 - 10.07) | <0.001 |
|  |  | pRBD with hyposmia vs controls  (105 vs 4243) | Group | 1.26 (1.04 - 1.54) | 0.021 |
|  |  |  | Age | 1.07 (1.03 - 1.11) | 0.004 |
|  |  |  | Sex (Male) | 3.50 (2.35 - 5.26) | <0.001 |
|  |  | Normosmic pRBD vs controls  (1201 vs 4243) | Group | 1.21 (1.14 - 1.30) | <0.001 |
|  |  |  | Age^2^ | 1.01 (1.01 - 1.01) | <0.001 |
|  |  |  | Sex (Male) | 2.65 (2.32 - 3.03) | <0.001 |
| DFT | KS | PSG-iRBD vs controls  (26 vs 1659) | Group | 1.07 (0.72 - 1.54) | 0.101 |
|  |  |  | Age^2^ | 1.01 (1.00 - 1.02) | 0.007 |
|  |  |  | Sex (Male) | 2.11 (7.21 - 9.00) | <0.001 |
|  |  | pRBD with hyposmia vs controls  (85 vs 1659) | Group | 1.52 (1.24 – 1.85) | <0.001 |
|  |  |  | Age | 1.03 (0.99 - 1.08) | 0.145 |
|  |  |  | Sex (Male) | 4.21 (2.68 - 6.68) | <0.001 |
|  |  | Normosmic pRBD vs controls  (502 vs 1659) | Group | 1.21 (1.10 -1.34) | <0.001 |
|  |  |  | Age^2^ | 1.00 (1.00 - 1.01) | 0.001 |
|  |  |  | Sex (Male) | 2.76 (2.24 - 3.41) | <0.001 |
|  | AT | PSG-iRBD vs controls  (26 vs 1659) | Group | 1.17 (0.75 -1.77) | 0.482 |
|  |  |  | Age^2^ | 1.01 (1.00 - 1.02) | 0.006 |
|  |  |  | Sex (Male) | 2.21 (7.45 - 9.47) | <0.001 |
|  |  | pRBD with hyposmia vs controls  (85 vs 1659) | Group | 1.64 (1.33 – 2.03) | <0.001 |
|  |  |  | Age | 1.03 (0.98 - 1.07) | 0.261 |
|  |  |  | Sex (Male) | 4.53 (2.86 -  7.26) | <0.001 |
|  |  | Normosmic pRBD vs controls  (502 vs 1659) | Group | 1.21 (1.10 -1.34) | <0.001 |
|  |  |  | Age^2^ | 1.00 (1.00 - 1.01) | <0.001 |
|  |  |  | Sex (Male) | 2.80 (2.27 - 3.46) | <0.001 |
|  | IS | PSG-iRBD vs controls  (26 vs 1659) | Group | 1.37 (0.93 - 1.98) | 0.101 |
|  |  |  | Age^2^ | 1.01 (1.00 - 1.02) | 0.007 |
|  |  |  | Sex (Male) | 2.08 (7.17 - 8.86) | <0.001 |
|  |  | pRBD with hyposmia vs controls  (85 vs 1659) | Group | 1.38 (1.12 - 1.70) | 0.002 |
|  |  |  | Age | 1.04 (1.00 - 1.08) | 0.071 |
|  |  |  | Sex (Male) | 3.63 (1.12 - 1.70) | <0.001 |
|  |  | Normosmic pRBD vs controls  (502 vs 1659) | Group | 1.24 (1.12 - 1.37) | <0.001 |
|  |  |  | Age^2^ | 1.00 (1.00 - 1.02) | <0.001 |
|  |  |  | Sex (Male) | 2.63 (2.14 - 3.23) | <0.001 |
| TMT | Executive function | PSG-iRBD vs controls  (99 vs 1601) | Group | 1.71 (1.40 -2.09) | <0.001 |
|  |  |  | Age^2^ | 1.01 (1.00 - 1.01) | <0.001 |
|  |  |  | Sex (Male) | 7.08 (4.37 - 11.19) | <0.001 |
|  |  | pRBD with hyposmia vs controls  (104 vs 1601) | Group | 1.16 (0.95 - 1.40) | 0.141 |
|  |  |  | Age | 1.06 (1.02 - 1.10) | 0.003 |
|  |  |  | Sex (Male) | 3.43 (2.28 - 5.21) | <0.001 |
|  |  | Normosmic pRBD vs controls  (1266 vs 1601) | Group | 1.35 (1.24 - 1.46) | <0.001 |
|  |  |  | Age^2^ | 1.01 (1.00 - 1.01) | <0.001 |
|  |  |  | Sex (Male) | 2.72 (2.32 - 3.18) | <0.001 |
|  | TMT-A reaction time | PSG-iRBD vs controls  (99 vs 1601) | Group | 1.77 (1.46 - 2.15) | <0.001 |
|  |  |  | Age^2^ | 1.01 (1.00 - 1.01) | <0.001 |
|  |  |  | Sex (Male) | 6.90 (4.30 - 11.14) | <0.001 |
|  |  | pRBD with hyposmia vs controls  (104 vs 1601) | Group | 1.63 (1.36 - 1.97) | <0.001 |
|  |  |  | Age | 1.04 (1.00 - 1.08) | 0.054 |
|  |  |  | Sex (Male) | 3.56 (2.36 - 5.43) | <0.001 |
|  |  | Normosmic pRBD vs controls  (1266 vs 1601) | Group | 1.24 (1.14 - 1.34) | <0.001 |
|  |  |  | Age^2^ | 1.01 (1.00 - 1.01) | <0.001 |
|  |  |  | Sex (Male) | 2.75 (2.35 - 3.21) | <0.001 |
|  | TMT-B reaction time | PSG-iRBD vs controls  (99 vs 1601) | Group | 2.29 (1.87 - 2.82) | <0.001 |
|  |  |  | Age^2^ | 1.01 (1.00 - 1.01) | <0.001 |
|  |  |  | Sex (Male) | 7.08 (4.37 - 11.19) | <0.001 |
|  |  | pRBD with hyposmia vs controls  (104 vs 1601) | Group | 1.57 (1.30 - 1.89) | <0.001 |
|  |  |  | Age | 1.04 (1.00 - 1.08) | 0.083 |
|  |  |  | Sex (Male) | 3.43 (2.28 - 5.23) | <0.001 |
|  |  | Normosmic pRBD vs controls  (1266 vs 1601) | Group | 1.48 (1.36 - 1.61) | <0.001 |
|  |  |  | Age^2^ | 1.01 (1.00 - 1.01) | <0.001 |
|  |  |  | Sex (Male) | 2.71 (2.32 - 3.18) | <0.001 |

Controls were used as the reference group. Odds ratios (OR) with 95% confidence intervals (CI) are shown for group comparisons adjusted for age and sex.

pRBD - probable REM sleep behaviour disorder, PSG-iRBD - polysomnography confirmed isolated REM sleep behaviour disorder, IS - incoordination score, AT - akinesia time, KS - kinesia score, BRAIN test - Bradykinesia Akinesia Incoordination test, DFT - Distal Finger Tapping Test, TMT - Trail Making Test.

**Supplementary table S4: Mean change in motor and cognitive scores over time (LME baseline vs followup analysis)**

|  | Control | Normosmic pRBD | pRBD with hyposmia | PSG-iRBD |
| --- | --- | --- | --- | --- |
| BRAIN test Change (mean ± SD / median ± IQR) | | | | |
| KS | 1.13 ± 13.53 | 1.72 ± 14.52 | 2.81 ± 16.06 | 0.60 ± 8.03 |
| AT | -1.63 ± 25.17 | -4.10 ± 36.15 | -3.85 ± 33.79 | 0.71 ± 32.90 |
| IS [log-transformed] | -0.28 ± 1.64 | -0.44 ± 1.72 | -0.56 ± 1.53 | -0.19 ± 1.23 |
| DFT Change (mean ± SD / median ± IQR) | | | | |
| KS | 3.06 ± 27.42 | 0.74 ± 28.44 | -0.64 ± 32.10 | -4.31 ± 12.73 |
| AT | -0.56 ± 26.82 | -0.98 ± 31.47 | -4.17 ± 23.64 | 14.64 ± 39.86 |
| IS [log-transformed] | 0.00 ± 1.16 | 0.06 ± 1.19 | 0.01 ± 1.04 | -0.14 ± 1.18 |
| TMT Change (mean ± SD) | | | | |
| Executive function | -1.67 ± 593.37 | -53.39 ± 574.14 | 87.79 ± 589.24 | 347.11 ± 1015.57 |
| TMT-A reaction time | -24.89 ± 360.16 | -0.32 ± 350.38 | 69.03 ± 453.05 | -65.31 ± 497.76 |
| TMT-B reaction time | -26.56 ± 540.76 | -53.70 ± 546.40 | 18.76 ± 634.87 | 281.81 ± 1054.43 |

Values are presented as mean ± SD for normally distributed variables and median with IQR for non-normally distributed variables. Measures are derived from the BRAIN test, DFT, and TMT. Groups include healthy controls, normosmic pRBD, pRBD with hyposmia, and PSG-iRBD.

pRBD - probable REM sleep behaviour disorder, PSG-iRBD - polysomnography confirmed isolated REM sleep behaviour disorder, IS - incoordination score, AT - akinesia time, KS - kinesia score, BRAIN test - Bradykinesia Akinesia Incoordination test, DFT - Distal Finger Tapping Test, TMT - Trail Making Test.

**Supplementary Table S5 — Longitudinal mixed-effects model results**

| Test | Outcome | Normosmic pRBD vs controls β (95% CI) | p value | pRBD with hyposmia vs controls β (95% CI) | p value | PSG-iRBD vs controls β (95% CI) | p value |
| --- | --- | --- | --- | --- | --- | --- | --- |
| BRAIN test | KS | -0.48 (-1.13 – 0.17) | 0.148 | 0.99 (-0.52 – 2.50) | 0.199 | -0.01 (-2.13 – 2.12) | 0.996 |
|  | AT | -0.21 (-2.66 – 2.25) | 0.870 | 3.44 (-0.74 – 7.17) | 0.070 | 3.01 (-2.11 – 8.13) | 0.250 |
|  | IS | -0.06 (-0.16 – 0.04) | 0.257 | 0.08 (-0.09 – 0.24) | 0.350 | -0.14 (-0.48 – 0.21) | 0.433 |
| DFT | KS | -1.83 (-7.35 – 3.69) | 0.515 | 7.52 (-5.78 – 20.8) | 0.267 | -0.22 (-20.5 – 20.0) | 0.983 |
|  | AT | -0.80 (-10.7 – 9.14) | 0.874 | 14.0 (-10.0 – 38.0) | 0.253 | -19.4 (-56.8 – 18.0) | 0.310 |
|  | IS | -0.05 (-0.193 – 0.087) | 0.456 | -0.59 (-1.31 – 0.13) | 0.108 | -0.25 (-1.39 – 0.89) | 0.664 |
| TMT | Executive function | -7.79 (-32.7 – 17.3) | 0.546 | 18.5 (-34.9 – 71.8) | 0.497 | -130.0 (-203.0 – -56.3) | 0.001 |
|  | TMT-A reaction time | -0.02 (-0.10 – 0.07) | 0.694 | 0.03 (-0.17 – 0.22) | 0.792 | -0.10 (-0.37 – 0.16) | 0.451 |
|  | TMT-B reaction time | 0.01 (-0.08 – 0.09) | 0.823 | -0.04 (-0.24 – 0.16) | 0.698 | 0.32 (0.05 – 0.59) | 0.020 |

Longitudinal changes in digital motor and cognitive performance from linear mixed-effects models. Results from linear mixed-effects models showing group-by-time interaction coefficients (β) with 95% confidence intervals (CI) comparing normosmic pRBD, pRBD with hyposmia and PSG-iRBD groups with controls. Models were adjusted for age and sex. Outcomes are derived from the BRAIN test, DFT, and TMT.

pRBD - probable REM sleep behaviour disorder, PSG-iRBD - polysomnography confirmed isolated REM sleep behaviour disorder, IS - incoordination score, AT - akinesia time, KS - kinesia score, BRAIN test - Bradykinesia Akinesia Incoordination test, DFT - Distal Finger Tapping Test, TMT - Trail Making Test.

**Supplementary Figure S1 — Baseline performance by test and group**

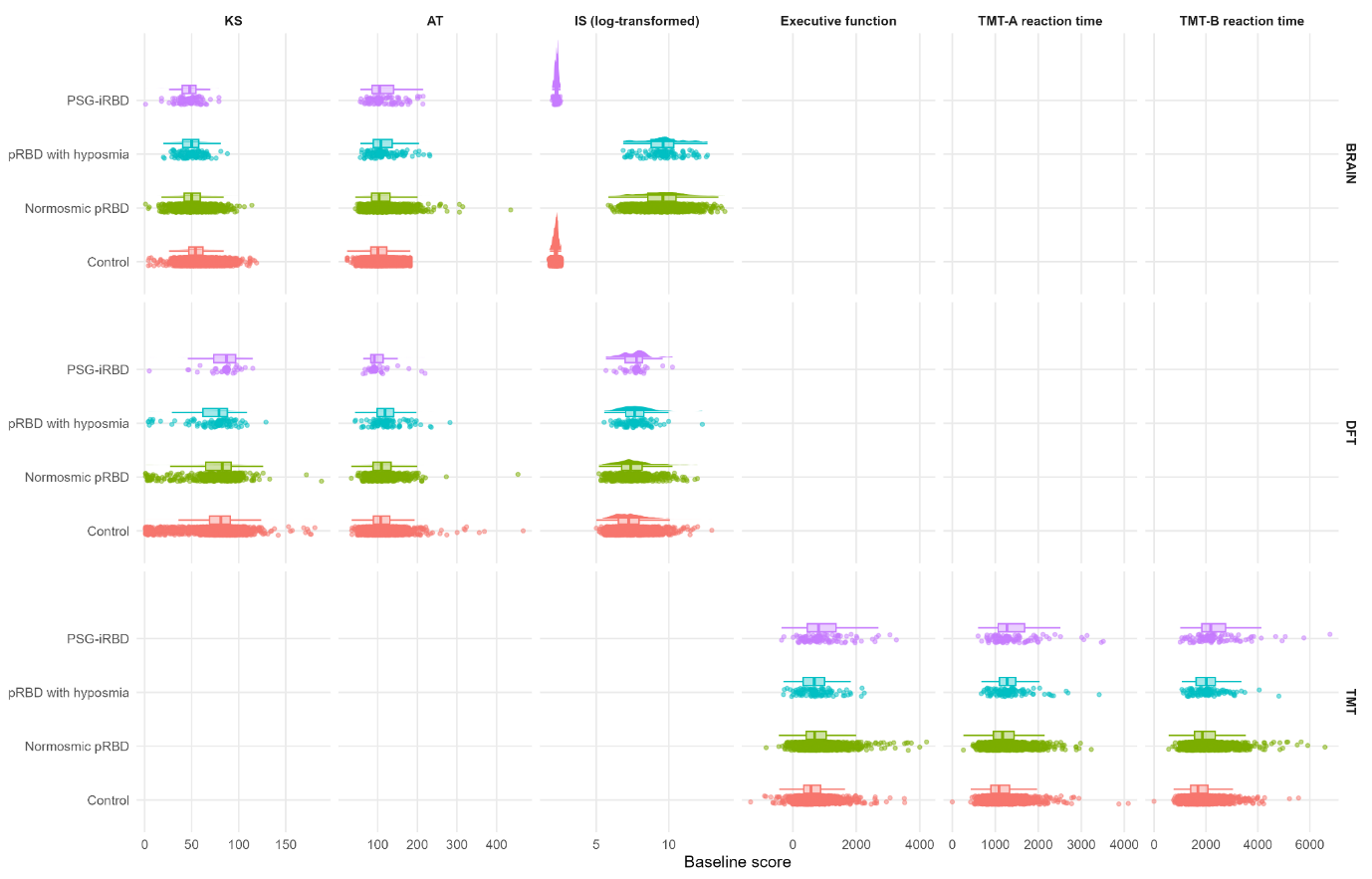

Raincloud plots showing the distribution of baseline scores for BRAIN, DFT, and TMT outcomes across controls, pRBD, pRBD with hyposmia, and PSG-iRBD. Points represent individual participants, boxplots show medians and interquartile ranges, and half-violins depict score distributions. IS values are log-transformed.

pRBD - probable REM sleep behaviour disorder, PSG-iRBD - polysomnography confirmed isolated REM sleep behaviour disorder, IS - incoordination score, AT - akinesia time, KS - kinesia score, BRAIN test - Bradykinesia Akinesia Incoordination test, DFT - Distal Finger Tapping Test, TMT - Trail Making Test.

**Supplementary Figure S2 — Mean trajectories over time (collapsed)**

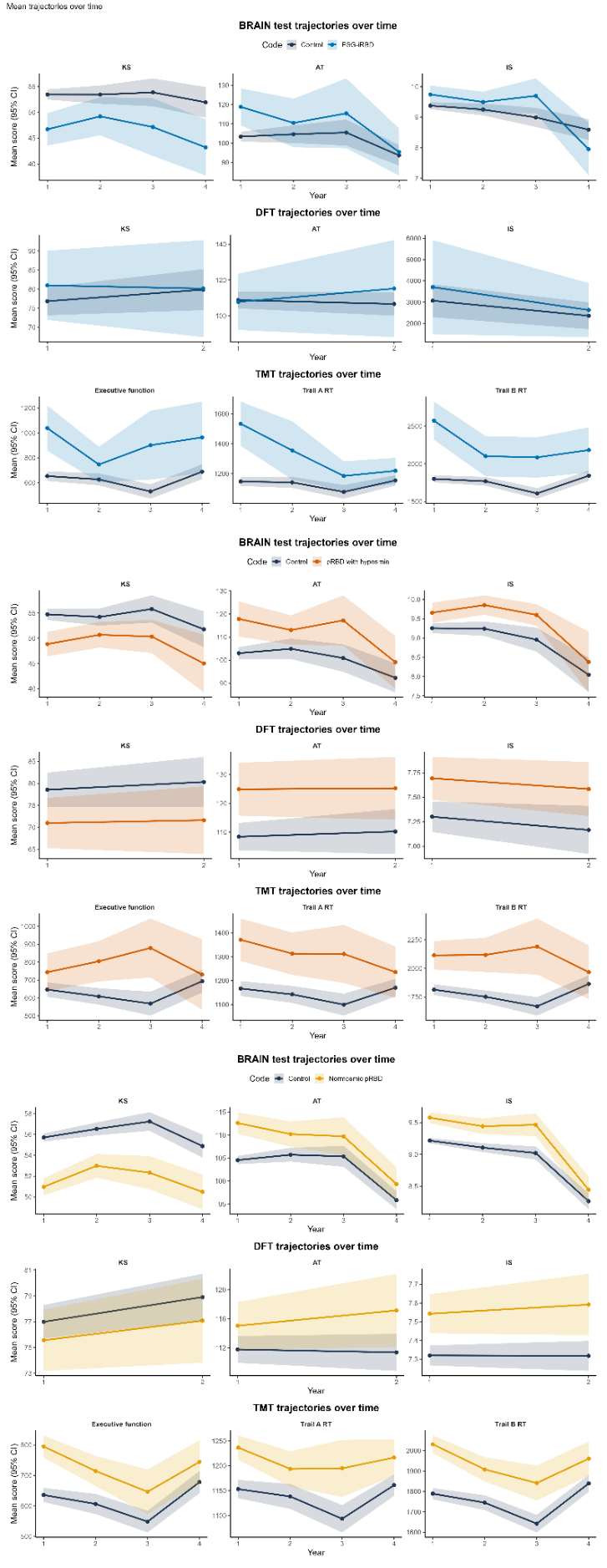

Longitudinal change in digital motor-cognitive performance in prodromal Parkinson's disease cohorts. Plots show mean performance across study visits for outcomes derived from the BRAIN test, DFT, and TMT. Lines represent group means and shaded areas represent 95% confidence intervals. Comparisons are presented between controls and RBD groups, including PSG-iRBD, pRBD with hyposmia, and normosmic pRBD. These trajectories illustrate differences in the rate of change in motor and cognitive performance across prodromal PD groups.

pRBD - probable REM sleep behaviour disorder, PSG-iRBD - polysomnography confirmed isolated REM sleep behaviour disorder, IS - incoordination score, AT - akinesia time, KS - kinesia score, BRAIN test - Bradykinesia Akinesia Incoordination test, DFT - Distal Finger Tapping Test, TMT - Trail Making Test.

**Supplementary Figure S3 — Heteroscedastic model diagnostics**

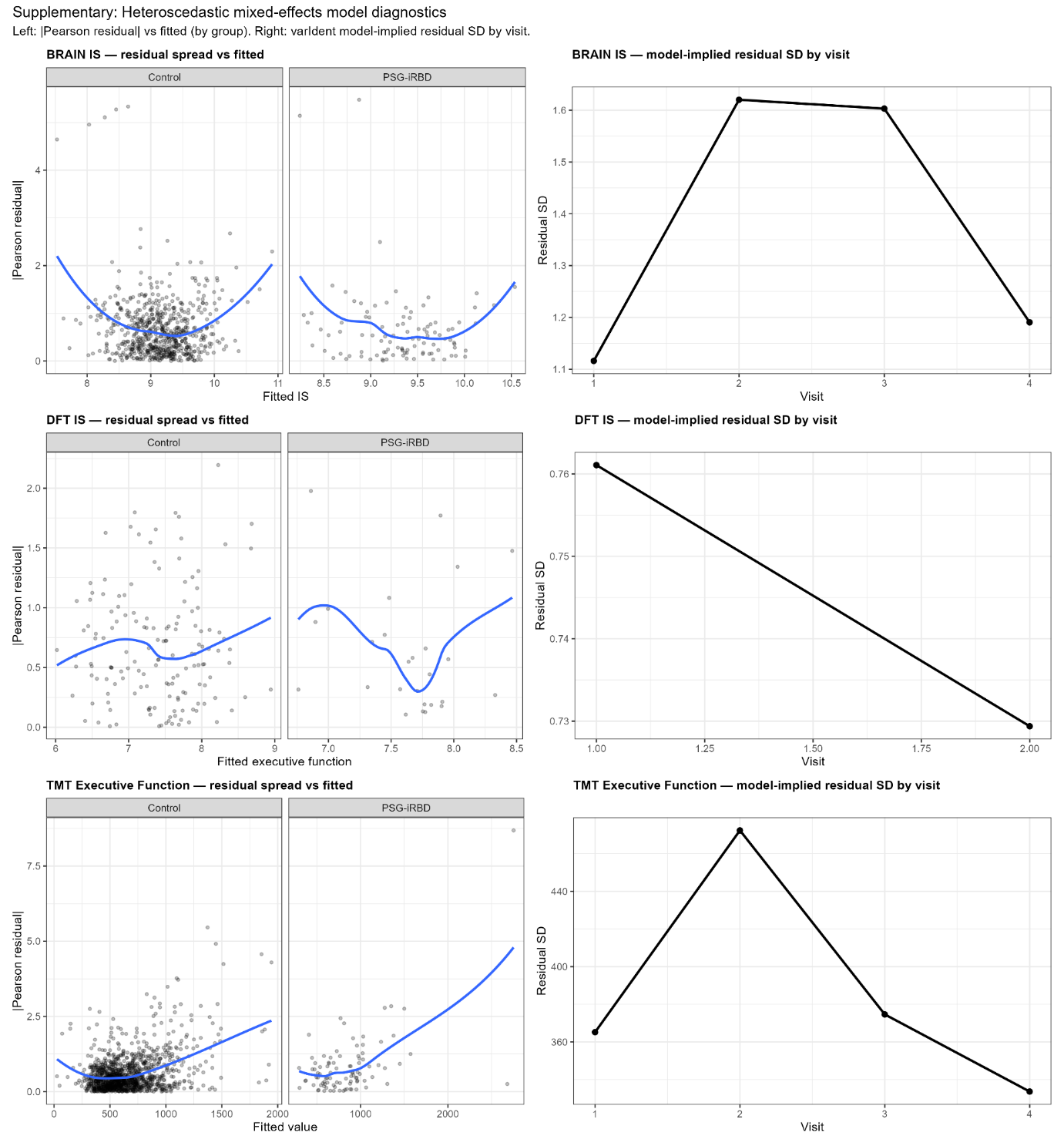

Left panels show Pearson residuals versus fitted values (stratified by group) for BRAIN test IS, DFT IS, and TMT executive function, illustrating residual spread across the fitted range. Right panels show model-implied residual standard deviations by visit from heteroscedastic mixed-effects models using visit-specific variance structures (varIdent). These diagnostics were used to assess time-varying variance in longitudinal models.

IS - incoordination score, BRAIN test - Bradykinesia Akinesia Incoordination test, DFT - Distal Finger Tapping Test, TMT - Trail Making Test.
